# Rainfall, Mosquito Indices, and Dengue Outbreaks in Southern Taiwan: Reassessing Predictive Modeling with Machine Learning Approaches

**DOI:** 10.1101/2025.09.10.25335502

**Authors:** Hsiang Hong

## Abstract

Dengue remains a major public health challenge in southern Taiwan, where recurrent outbreaks are shaped by complex environmental and entomological drivers. Using nine years of surveillance data (2015–2024) comprising more than 70,000 dengue cases, daily rainfall, and six mosquito indices from Tainan, Kaohsiung, and Pingtung, we evaluated the predictive value of these indicators through lagged correlation and machine learning models. Rainfall showed little explanatory power, while entomological indices demonstrated only moderate, location-specific associations, with the Container Index in Kaohsiung peaking at r ≈ 0.38 at 4–6 week lags. Regression models failed to predict outbreak magnitude, with R^2^ near zero on test data, but classification frameworks distinguishing outbreak from non-outbreak weeks achieved high accuracy (AUC up to 0.98), driven primarily by mosquito indices. These findings highlight the limited utility of regression forecasts for dengue, while supporting classification-based approaches as more practical tools for early outbreak detection and vector control planning.

## Introduction

Dengue fever is a rapidly expanding vector-borne disease in tropical and subtropical regions, posing significant public health challenges worldwide^1–3^. In Taiwan, the southern counties of Tainan, Kaohsiung, and Pingtung consistently report the highest dengue burden, with recurring outbreaks exacerbated by complex environmental and entomological drivers^4,5^. Climatic factors, especially precipitation and temperature, are known to influence *Aedes* mosquito breeding and survival, creating a temporal window for increased transmission risk^6–8^. Entomological surveillance indices such as Container Index (CI), Breteau Index (BI), and Pupal Index are widely used for early detection of vector proliferation, yet their effectiveness in predicting human cases remains debated^9–11^.

Several studies have employed machine learning (ML) techniques to forecast dengue incidence, often achieving high accuracy using meteorological and entomological predictors in retrospective analyses^12–14^. Notably, Kuo et al. (2024) obtained robust predictive performance (AUC ≈ 0.95) by training models on short-term (2013–2015) data splits, relying on rainfall, air quality, and UV indices^15^. However, model performance in such in-sample validations may overestimate generality, especially when future outbreaks are unprecedented in magnitude^16^.

In this study, we address two critical gaps. First, we systematically evaluate the temporal relationships between rainfall, six entomological indices (CI, AI, BI, Containers per 100 households, House Index for *Aedes aegypti*, and Pupal Index), and dengue incidence across three southern Taiwanese counties from 2015 to early 2024. Second, we assess predictive modeling under two frameworks: regression to estimate case counts, and classification to distinguish outbreak versus non-outbreak weeks. By explicitly demonstrating the limitations of regression for forecasting rare outbreak magnitudes and highlighting the utility of ML classification for outbreak detection, we provide a more nuanced and realistic appraisal of prediction tools for dengue surveillance.

## Results

### Data Collection

To investigate meteorological and entomological influences on dengue incidence in Taiwan, we compiled surveillance and environmental data from December 28, 2015 to December 30, 2024, covering a total of 9,863 daily observations. During this period, more than 70,000 laboratory-confirmed dengue cases were reported nationwide, of which we focused on the southern counties of Tainan, Kaohsiung, and Pingtung, the regions with the highest disease burden.

Daily dengue case records, mosquito surveillance indices, and precipitation measurements were aggregated to the weekly level to align temporal resolution across datasets and to account for the time-lagged effects of climate and entomological drivers. This resulted in a dataset comprising 1,409 county-weeks of aligned observations, with variables including weekly case counts, cumulative precipitation (mm), and six entomological indices (Container Index [CI], Adult Index [AI], Breteau Index [BI], Containers per 100 households, House Index for *Aedes aegypti*, and Pupal Index).

### Temporal Dynamics of Dengue, Rainfall, and Mosquito Indices

Having assembled and harmonized dengue case counts, rainfall, and mosquito surveillance indices into a common weekly framework, we next examined the temporal dynamics of these variables across the three high-burden counties of Tainan, Kaohsiung, and Pingtung (Fig. 1a-c).

**Figure 1.**
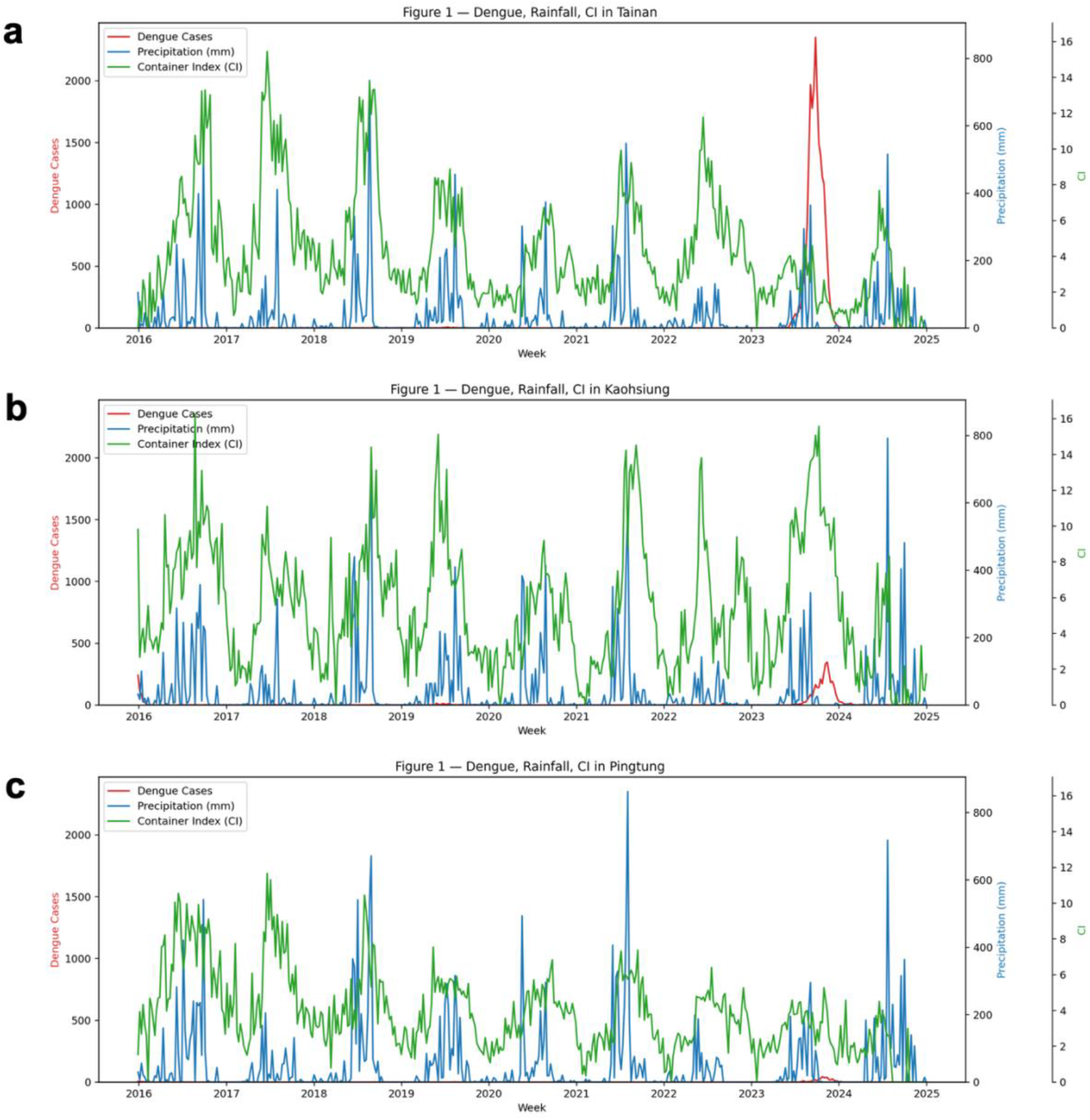
Temporal dynamics of dengue incidence, rainfall, and mosquito indices in southern Taiwan (2016–2024). (a) Tainan, (b) Kaohsiung, and (c) Pingtung. Weekly dengue cases (red, left y-axis), accumulated weekly rainfall (blue, right y-axis), and mosquito Container Index (CI; green, secondary right y-axis) are shown. Peaks in rainfall consistently occur during the summer monsoon, while CI fluctuates seasonally, reflecting *Aedes aegypti* breeding conditions. Large dengue outbreaks were observed in Tainan (2015–2016 and 2023–2024) and in Kaohsiung (2023–2024), whereas Pingtung reported relatively fewer cases. While increases in CI often followed rainfall peaks, their correspondence with dengue incidence was inconsistent, underscoring the multifactorial drivers of epidemic onset.

From 2015 through 2023, dengue incidence in southern Taiwan displayed marked temporal and geographic heterogeneity. Tainan experienced the most severe outbreaks, including the nationwide epidemic of 2015 and another large resurgence in 2023, with weekly peaks exceeding 2,000 reported cases. Kaohsiung exhibited intermediate activity, with outbreaks reaching several hundred weekly cases, whereas Pingtung reported relatively lower case counts throughout the study period. These patterns reaffirm the disproportionate burden borne by the southernmost counties of Taiwan^5^.

Rainfall followed a regular seasonal cycle, peaking during the summer monsoon months, while mosquito surveillance indices fluctuated within and across years. Although periods of elevated precipitation often coincided with increased values of indices such as the Container Index (CI), these trends did not consistently align with epidemic onset. Notably, elevated CI values in Kaohsiung were observed to precede local outbreaks, whereas similar increases in Tainan and Pingtung were not always accompanied by a rise in dengue incidence. This suggests that meteorological and entomological drivers provide only partial signals for outbreak potential, with other factors such as viral introduction, human mobility, and population immunity likely contributing to the timing and magnitude of epidemics^10,11,17^.

### Lagged Correlations: Rainfall and Mosquito Indices

To assess whether environmental or entomological indicators precede dengue activity, we next examined lagged correlations between weekly dengue cases and rainfall or mosquito indices across the three southern counties.

Across all three counties, rainfall demonstrated weak correlations with dengue incidence (r ≤ 0.1 in most cases). In Tainan and Pingtung, rainfall showed small positive associations at lags of 3–8 weeks, consistent with the expected delay between precipitation, mosquito breeding, and subsequent virus transmission, though the signal remained marginal. In Kaohsiung, rainfall correlations were negligible across all lags.

Mosquito indices demonstrated similarly mixed results. Among the six entomological indices evaluated (Container Index [CI], House Index [HI], Breteau Index [BI], Con100HH, HI*Aeg, and pupal counts), the CI in Kaohsiung achieved the strongest association, peaking at r ≈ 0.38 with dengue cases at a lag of 4–6 weeks (Fig. 2a–c). However, in Tainan and Pingtung, none of the indices showed consistent or meaningful correlations, with most values fluctuating near zero or negative.

**Figure 2.**
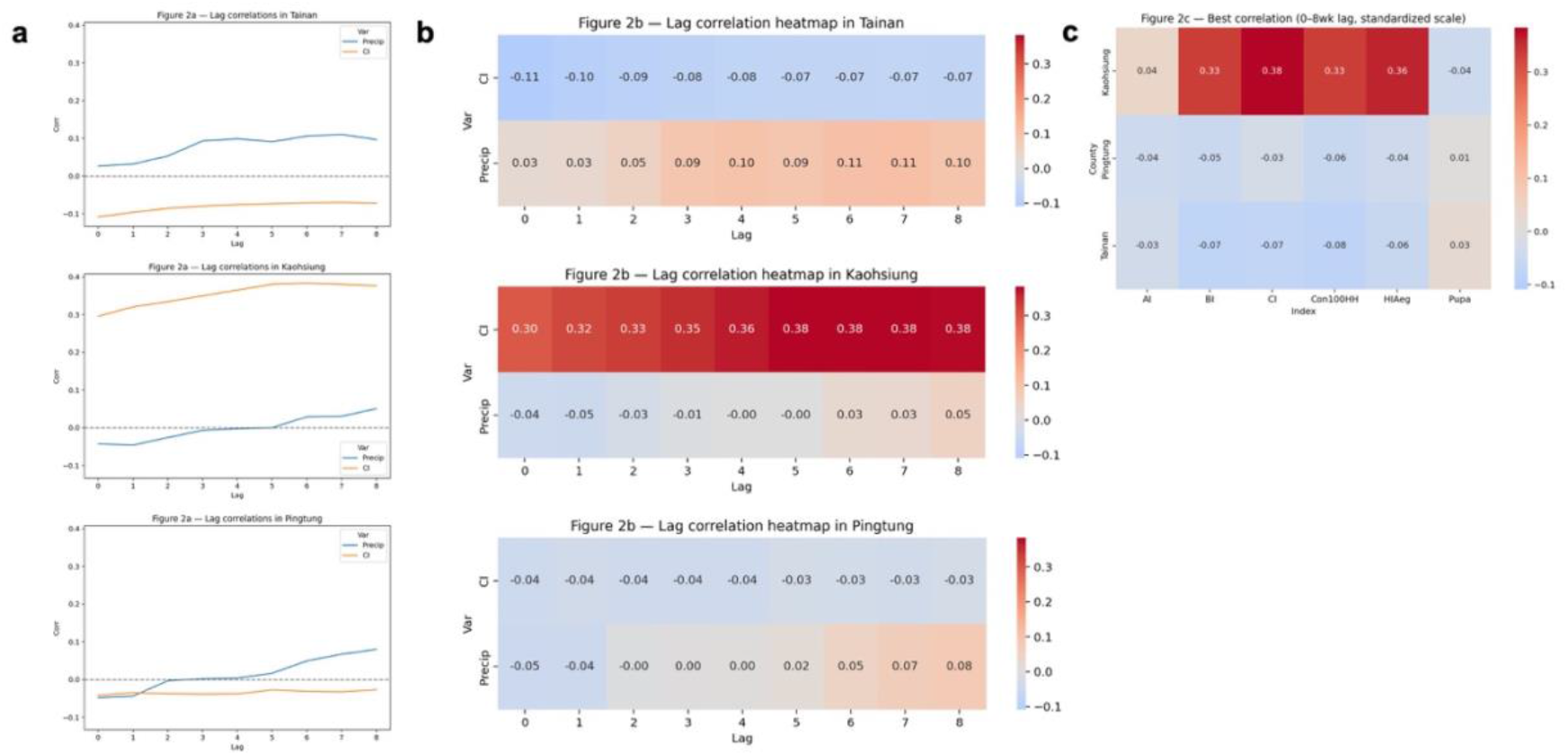
Lagged correlations between dengue incidence and environmental predictors in southern Taiwan. (a) Line plots of Pearson correlations (r) between weekly dengue cases and precipitation (blue) or Container Index (CI; orange) at 0–8 week lags, shown separately for Tainan, Kaohsiung, and Pingtung. Kaohsiung demonstrated the strongest associations, with CI rising to r ≈ 0.38 at 5–6 week lags, while correlations in Tainan and Pingtung remained weak. (b) Heatmaps of lagged correlations for precipitation and CI across 0–8 week lags in each county. (c) Heatmap of the best correlation (maximum across 0–8 week lags) for all six mosquito indices (AI, BI, CI, Con100HH, HIAeg, Pupa). Moderate correlations (r ≈ 0.3– 0.4) were observed only for Kaohsiung (CI, HIAeg, Con100HH), while other indices and regions yielded negligible associations.

These results highlight two key points: (i) traditional entomological indices do not reliably predict dengue activity in all settings, and (ii) correlations are highly location-dependent, with Kaohsiung showing moderate association but Tainan and Pingtung exhibiting weak or absent relationships. This underscores the challenge of using single predictors in isolation to anticipate outbreaks and suggests that integrative, multi-factorial models may be necessary.

### Regression Models Fail to Predict Outbreak Magnitude

Given the weak and inconsistent associations observed when rainfall and mosquito indices were evaluated individually, we next asked whether combining multiple predictors within a machine learning framework could enhance forecasting performance. Specifically, we tested both regression approaches (predicting weekly dengue case counts directly) and classification approaches (predicting whether a given week would be an outbreak or non-outbreak week).

For regression, we implemented three widely used ensemble methods, Random Forest, Gradient Boosting, and XGBoost, using rainfall and mosquito indices as lagged predictors (Fig. 3). However, all three regressors failed to meaningfully capture outbreak magnitude, producing near-zero or negative explanatory power (R^2^ ≈ –0.05 across models; RMSE ≈ 336). Instead of tracking the epidemic peaks, predictions collapsed toward the baseline, underestimating case counts during outbreak weeks and over-smoothing during inter-epidemic periods.

**Figure 3.**
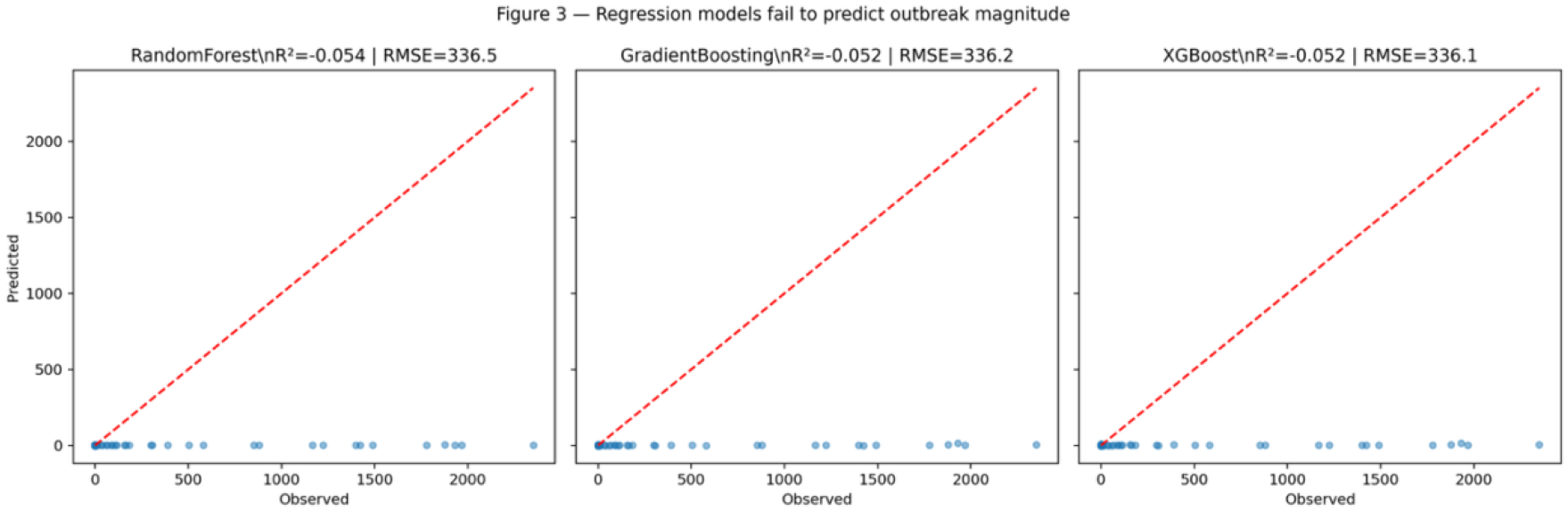
Regression models fail to predict outbreak magnitude. Scatterplots comparing observed versus predicted weekly dengue cases in southern Taiwan using Random Forest, Gradient Boosting, and XGBoost regressors. Despite incorporating rainfall and mosquito indices as predictors, all models performed poorly (R^2^ < 0, RMSE ≈ 336), with predictions clustered near zero and failing to capture epidemic peaks. The dashed red line indicates the 1:1 line of perfect prediction.

This consistent failure underscores a key limitation of regression-based dengue forecasting: the statistical distribution of dengue incidence is highly imbalanced, with long stretches of zero or near-zero cases punctuated by sudden epidemic surges. Standard regression frameworks, which assume relatively stable variance, tend to regress toward the mean, rendering them poorly suited to predicting outbreak magnitudes. Importantly, this limitation was not alleviated by the choice of algorithm — tree-based methods that often excel in nonlinear relationships performed no better than gradient boosting approaches optimized for predictive accuracy.

Taken together, these results demonstrate that simply adding multiple meteorological and entomological predictors into a regression model is insufficient for outbreak forecasting. Rather than capturing outbreak severity, the models defaulted to minimizing error during the much more frequent non-outbreak periods. Instead, it may be more effective to reframe the problem as a classification task (outbreak vs. non-outbreak weeks), where the skewed distribution of dengue incidence can be handled more appropriately and where categorical early-warning signals are more actionable for public health planning.

### Classification Models Distinguish Outbreak from Non-outbreak Weeks

While regression approaches were unable to capture outbreak magnitudes, re-framing the problem as a binary classification task (outbreak vs. non-outbreak weeks) yielded substantially better performance. We trained logistic regression, Random Forest, Gradient Boosting, and XGBoost classifiers on lagged rainfall and mosquito indices, defining outbreak weeks as those exceeding the 90th percentile of weekly dengue cases.

Receiver operating characteristic (ROC) curves demonstrated that tree-based ensemble methods strongly outperformed logistic regression (Fig. 4a). Random Forest achieved the highest discriminatory power (AUC = 0.98), followed by XGBoost (AUC = 0.93) and Gradient Boosting (AUC = 0.92), while logistic regression showed only moderate performance (AUC = 0.77).

**Figure 4.**
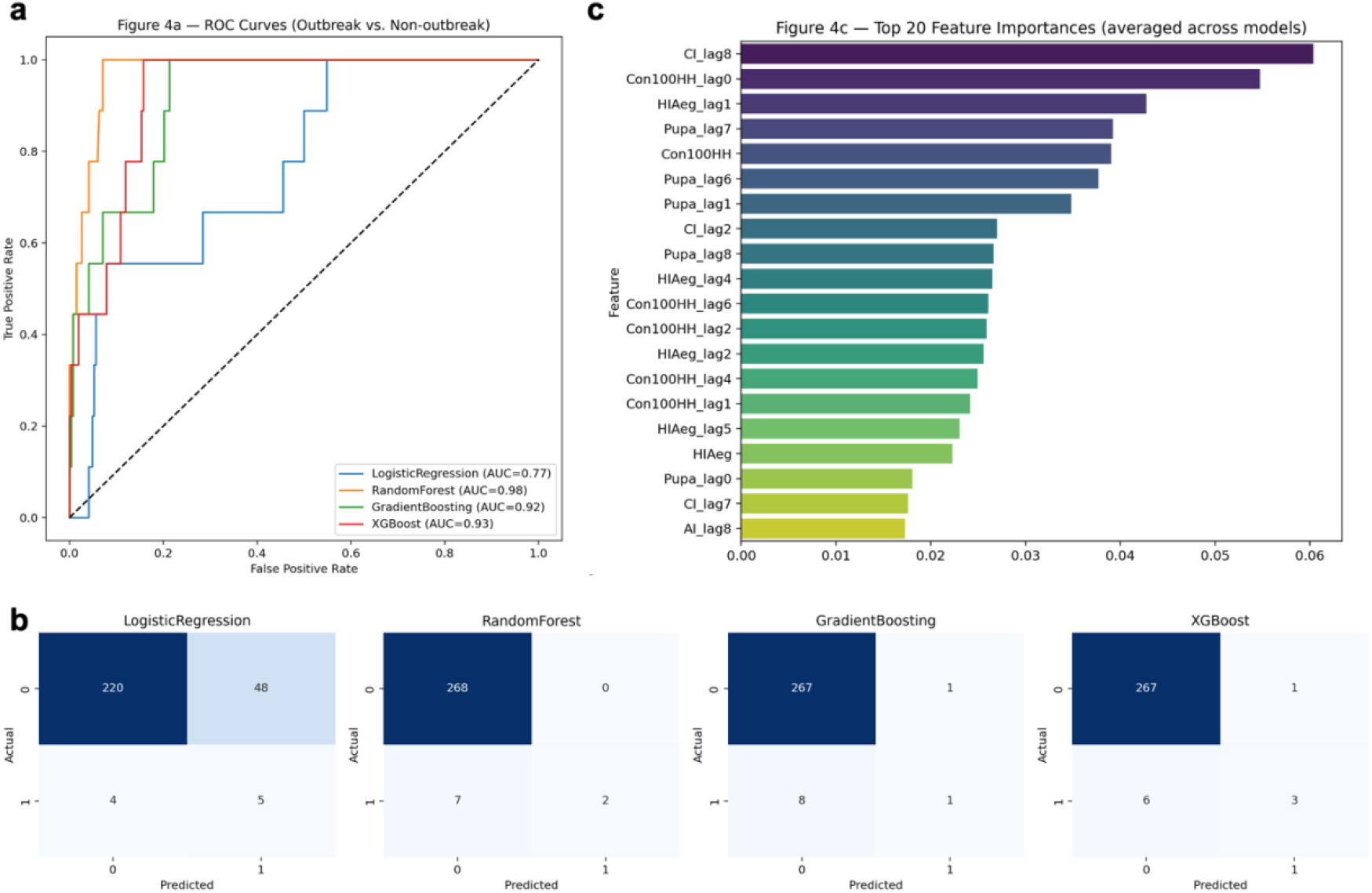
Classification performance for outbreak detection in southern Taiwan. (a) Receiver operating characteristic (ROC) curves for logistic regression, Random Forest, Gradient Boosting, and XGBoost classifiers distinguishing outbreak vs. non-outbreak weeks. Ensemble methods substantially outperformed logistic regression, with Random Forest achieving the highest AUC (0.98). (b) Confusion matrices of observed vs. predicted outbreak status on the held-out test set. Logistic regression misclassified nearly half of outbreak weeks, while tree-based models correctly identified most outbreak periods. (c) Feature importance rankings (averaged across ensemble models), highlighting lagged entomological indices (CI, HIAeg, Con100HH, pupal counts) as the strongest predictors, while rainfall variables contributed little.

Confusion matrices further underscored these differences (Fig. 4b). Logistic regression misclassified nearly half of outbreak weeks, whereas ensemble classifiers correctly identified the majority, albeit at the expense of some false positives. Random Forest, in particular, achieved nearly perfect separation of outbreak and non-outbreak weeks in the test set.

Feature importance analysis indicated that lagged mosquito indices, especially container index (CI), HI*Aeg, and Con100HH, were the strongest drivers of classification (Fig. 4c). Pupal counts also contributed meaningfully, while rainfall lagged by up to 8 weeks showed lower and less consistent influence. This pattern suggests that entomological surveillance indicators may provide more immediate predictive value for outbreak detection than meteorological inputs alone.

## Discussion

This study evaluated the predictive value of rainfall and entomological indices for dengue outbreaks in southern Taiwan across a nine-year period. While exploratory correlation analyses revealed weak and inconsistent associations between individual predictors and dengue incidence, our machine learning framework demonstrated two critical insights: (i) regression models cannot reliably forecast outbreak magnitude, and (ii) classification approaches offer greater promise for distinguishing outbreak versus non-outbreak periods.

Our findings contrast with previous reports that achieved high predictive accuracy using regression-based approaches in Taiwan and other dengue-endemic setting^15,18,19^. We show that such results may reflect model overfitting within outbreak periods rather than generalizable predictive power across unseen data. By explicitly framing outbreak prediction as a classification task, we emphasize the non-linear and threshold-like nature of dengue transmission dynamics, where environmental and entomological conditions may be sufficient to tip systems into epidemic states but do not scale proportionally with case counts^20^.

Importantly, the most informative predictors identified by our models were lagged entomological indices — particularly CI, HIAeg, Con100HH, and pupal counts — while rainfall contributed little explanatory value. This finding highlights the primacy of vector surveillance for early outbreak detection and aligns with broader entomological research suggesting that vector abundance is a more direct driver of transmission than meteorological conditions alone^10,11,21^. Nonetheless, correlations varied substantially by location, with moderate associations observed in Kaohsiung but negligible signals in Tainan and Pingtung. This heterogeneity underscores the challenge of extrapolating predictive models across regions and supports the need for localized calibration of early warning systems^6,7^.

Several limitations should be noted. Our models relied only on rainfall and six standard mosquito indices, without incorporating other potentially important predictors such as temperature, humidity, human mobility, or land-use factors^22–25^. Moreover, all analyses were restricted to data from Tainan, Kaohsiung, and Pingtung between 2015 and 2024, and we did not validate the models on external datasets, limiting the generalizability of our findings to other regions or time periods^6^.

Despite these limitations, our results provide an important corrective to overly optimistic regression-based forecasts^26^ and demonstrate the value of classification-oriented approaches. Outbreak detection models may serve as practical tools for public health decision-making, helping to prioritize intensified surveillance or vector control during high-risk periods^27^, even if precise case numbers cannot be forecasted. Future studies should extend this work by integrating additional data sources, testing hybrid mechanistic–statistical frameworks, and evaluating performance prospectively in real-world public health systems.

## Methods

### Data Collection

We compiled three datasets covering the period from December 28, 2015 to December 30, 2024: (i) daily dengue case counts from the Taiwan CDC’s National Notifiable Disease Surveillance System, (ii) daily rainfall records from the Central Weather Administration (CWA), and (iii) mosquito surveillance data from the Taiwan CDC entomological monitoring program. The latter included six indices: Container Index (CI), House Index (HI), Breteau Index (BI), Con100HH, HIAeg, and pupal counts. Data were restricted to the three southern counties of Tainan, Kaohsiung, and Pingtung, which consistently report the highest dengue burden in Taiwan.

### Preprocessing and aggregation

Daily records were harmonized by county through standardized mapping of administrative divisions. For precipitation, station metadata were used to infer county-level assignments. All data were aggregated to epidemiological weeks beginning on Monday. Weekly dengue case counts were summed, precipitation was aggregated as the total weekly rainfall (mm), and mosquito indices were averaged across stations within each county. The final dataset comprised 1,409 county-week observations with non-missing data across all three sources.

### Machine learning models

We evaluated both regression and classification frameworks. For regression, Random Forest, Gradient Boosting, and XGBoost were trained to predict weekly case counts from lagged rainfall and mosquito indices. Models were trained on the first 80% of weeks and tested on the final 20%. For classification, outbreak weeks were defined as those exceeding the 75th percentile of weekly case counts, and logistic regression, Random Forest, Gradient Boosting, and XGBoost classifiers were trained to distinguish outbreak versus non-outbreak weeks. Performance was assessed using R^2^ for regression, and area under the receiver operating characteristic curve (AUC), confusion matrices, and feature importance rankings for classification.

### Statistical analysis

All analyses were performed in Python (v3.11) on Google Colab using pandas, scikit-learn, xgboost, and matplotlib. Code for preprocessing, correlation analysis, and model training is available upon request.

## Data Availability

All data produced in the present study are available upon reasonable request to the authors

